# Significantly longer Covid-19 incubation times for the elderly, from a case study of 136 patients throughout China

**DOI:** 10.1101/2020.04.14.20065896

**Authors:** Ally Bi-Zhu Jiang, Richard Lieu, Siobhan Quenby

**Affiliations:** Shenzhen RAK wireless Technology Co., Ltd., China; Department of Physics, University of Alabama, Huntsville, AL 35899, USA; Division of Reproductive Health, Warwick Medical School, The University of Warwick, UK

## Abstract

**Objective:** To infer Covid-19 incubation time distribution from a large sample.

**Method:** Based on individual case data published online by 21 cities of China, we investigated a total of 136 COVID-19 patients who traveled to Hubei from 21 cities of China between January 5 and January 31, 2020, remained there for 48 hours or less, and returned to these cities with onset of symptoms between January 10 and February 6, 2020. Among these patients, 110 were found to be aged 15 – 64, 22 aged 65 – 86, and 4 aged under 15.

**Findings:** The differential incubation time histogram of the two age groups 15 – 64 and 65 – 86 are adequately fitted by the log normal model. For the 15 - 64 age group, the median incubation time of 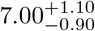 days (uncertainties are 95 −0.90 % CL) is broadly consistent with previous literature. For the 65-86 age group, the median is 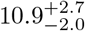 days is statistically significantly longer. Moreover, for −2.0 this group, the 95 % confidence contour indicates the data cannot constrain the upper bound of the log normal parameters *µ, σ* by failing to close there; this is because the sample has a maximum incubation time of 17 days, beyond which we ran out of data even though the histogram has not yet peaked. Thus there is the potential of a much longer incubation time for the 65-86 age group than 10 – 14 days. Only a much larger sample can settle this.

## 1. Introduction

The incubation time of Covid-19 [1] and the closely related question of asymptomatic case numbers are two topics of major interest and concern. On the former, the research results presented here for the main age group of 15-64 broadly corroborates previous studies [2, 3, 4, 5, 6], but for the elderly group of 65-86 years the incubation time we report below is significantly longer.

For nearly every city in China, daily information on list of COVID-19 cases is released officially to the Chinese social media WeChat accounts of respective cities. However, only a minority of cities include in their official release clear information on the day of symptom onset, which is required in estimating incubation period. We extracted the information so released between January 22 and February 15, 2020 and compiled a list of patients with COVID-19 who have traveled to Hubei, the origin of the COVID-19 pandemic, from cities which reported the day of symptom onset. The supplementary Excel file containing these raw data on COVID-19 cases reported from 21 cities of China outside Hubei: patient case number, age, sex, first and last day in Hubei, and first day with symptoms.

In the current investigation we included only those COVID-19 patients who stayed in Hubei for at most two calendar days. The day of exposure was taken as the first day to Hubei if the patient stayed in Hubei for one calendar day; or as the middle of the first and second day in Hubei if the patient stayed for two calendar days. By excluding COVID-19 patients who stayed in Hubei for more than two days, one can better define the the day of exposure. The incubation period for each COVID-19 patient is inferred as the number of days between exposure and symptom onset.

## 2. Log normal distribution

As will be shown below, the distribution of incubation times may adequately be fitted with a log normal distribution for the two age groups mentioned above, suggesting that the incubation time *τ* (in days) is a multiplicative variate. Specific to the problem of Covid-19, it seems reasonable to envisage an inverse proportionality relationship between the virus growth rate 1+r and *τ*, and the sample average growth rate is

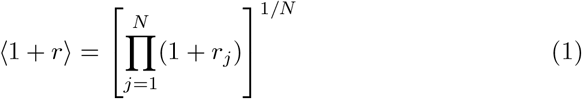

where *N* is the sample size (or number of measurements of the incubation time *τ*). Since *τ*_*j*_ ∝ 1*/*(1 + *r*_*j*_) this means

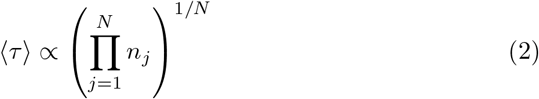

or, equivalently, both ln(1 + *r*) and ln *τ* are additive variates. Moreover, if ln(1 + *r*_*j*_) is normally distributed because 1 + *r*_*j*_ itself is a geometrically many times averaged growth rate of the virus inside the human body (*i*.*e*. the Central Limit Theorem may apply to ln(1 + *r*_*j*_)), the distribution of *τ*_*j*_ would then be a log normal^1^ of (arithmetic) mean *µ* and standard deviation *σ*. Thus the expected number of cases within some incubation time interval *k*, or incubation time *τ*_*k*_ = *kτ*_0_, is

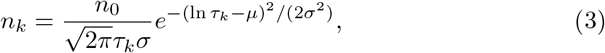

with the coefficient 1*/τ*_*k*_ originating from the relationship between logarithmic and linear intervals, *viz. d* ln *τ* = *dτ/τ*.

The applicability of the log normal model to the Covid-19 incubation times distribution compels one to calculate the mean incubation time as the geometric mean (2) at least as an alternative, as we shall do in the following section.

## 3. Fitting the data

The usual way of fitting a multi-parameter model to the data is by minimizing the *χ*^2^ statistic

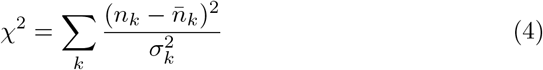

w.r.t. the model parameters *α, β, γ*, …, where *n*_*k*_ ≡ *n*_*k*_(*α, β, γ*, …) is the number of cases for incubation time interval *k* as expected by the model, 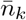 is the observed number of cases, and 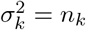 is the expected model variance assuming Poisson fluctuation in the case counts. In the case of the currently scrutinized log normal model distribution consisting of three parameters *α* = *µ, β* = *σ*, and *γ* = *n*_0_ (where *µ, σ*, and *n*_0_ are as in (3)), 95 % confidence intervals *δµ, δσ*, and *δn*_0_ for the best-fit parameters may be inferred from the Δ*χ*^2^ criterion discussed below. For a log normal quantile *q* = *q*(*m*) = *e*^*µ*+*mσ*^, Δ*χ*^2^ also yields *δq*. Yet an alternative method is to take advantage of the independence of *µ* and *σ* in the model by writing

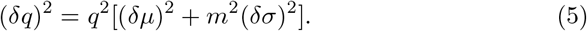

It turns out, however, that the fitting technique of (4) leads to large uncertainties in some parameters of interest, including those computed within the dynamic range of incubation times as set by the data. This is especially the case for the second of the two age groups, consisting of age 65-86 years patients, where the incubation time span of the model is significantly wider than the data. Below we present a slight variation of the method in (4) which avoids the problem.

To facilitate introducing the modified model fitting algorithm we first remind the reader of the standard maximum likelihood method^2^, which relies upon the *n*_*k*_ ≫ 1 limit for all time intervals *k*, the limit where a Poisson distribution of counts tend to a Gaussian with equality between variance and mean. Thus the likelihood of a match between the number of counts *n*_*k*_ predicted by the model and observational data 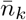 for *all* incubation time intervals *k* is given by the conditional probability

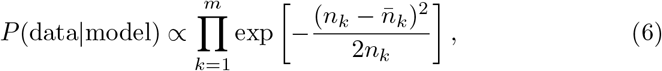

and one’s task is to maximize *P* (data|model) w.r.t. the model parameters *α, β, γ*, …, where *m* is the total number of time intervals spanned by the data. If the number of counts per bin does not satisfy *n*_*k*_ ≫ 1, however, the distribution of counts will not be normal, *i*.*e*. it will not be genuinely Poisson, in which case the expression on the right side of (6) is not strictly valid. In this limit, which does apply to the second of our two age groups (*viz*. 65 – 86 of age), one should use likelihood ratios [7] instead of (6), but because the difference between the two output parameter values are small compared to their uncertainties there is no real advantage in deviating from (6); we simply feel that for completeness sake this subtle point should be mentioned.

The maximization of (6) w.r.t. model parameters is obviously the same as minimizing *χ*^2^ as given in (4). Explicitly, if one writes, in the context of the log normal model (3),

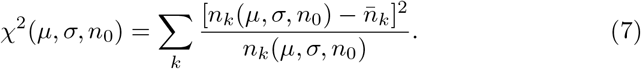

the procedure would be equivalent to solving

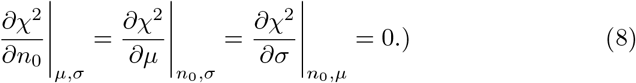

Thus there are 3 equations in 3 unknowns, and the minimization process is fully deterministic.

If, as mentioned above, the total incubation time *mτ*_0_ spanned by the entire database is not long enough to clinch the full extent of the log normal distribution, one will have to constrain the fitting procedure to ensure that the area under the log normal is *exactly* equal to the total number of cases *N* over the time *mτ*_0_. The specific question one seeks to answer here is: given there are *N* cases to be randomly distributed into *m* time intervals in accordance with a prescribed set of average proportions {*n*_*k*_}, *k* = 1, 2, …, *m* satisfying

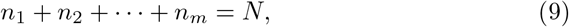

and *n*_*k*_ = *n*_*k*_(*n*_0_, *µ, σ*), how would one tune *n*_0_, *µ*, and *σ* to maximize the likelihood of the hypothetical distribution agreeing with the data, when Poisson counting uncertainties in the latter are taken into account? Note that the model for *n*_*k*_ does not have to cut off at *k* = *m, i*.*e*. (9) is merely there to enforce the equality between expected and actual total number of cases within the full range of incubation times available to the study. In this way, one is obliged to respect only those ‘in range’ parameter values ensuing from the best fit model.

Thus one would now extremize the statistic

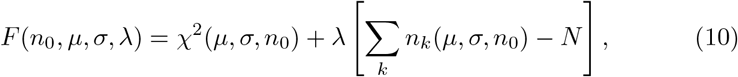

by requiring

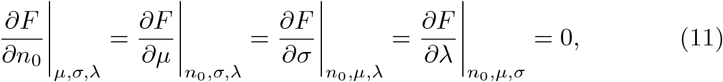

where the vanishing of the last partial derivative enforces the area (*λ* is a Lagrange multiplier). Once again there are 4 equations solving for 4 unknowns, and the number of free parameters is reduced from the previously 3 to currently 2 (note however that the area constraint is not as simple as fixing *n*_0_; note also that the degrees of freedom of the whole problem is increased from the *m* – 3 to *m* − 2).

Turning to the confidence interval for an interested parameter *η*(*n*_*k*_) = *η*(*µ, σ, n*_0_), such as the expected (arithmetic) mean incubation time within the observation interval *mτ*_0_,

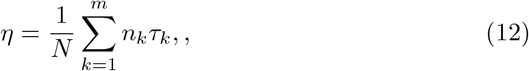

one could re-extremize *F* subject to yet another additional constraint which ensures *η* equals some fixed value *η*_0_ by invoking one more Lagrange multiplier *ν* to form the *G* statistic,

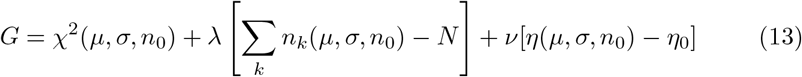

and requiring

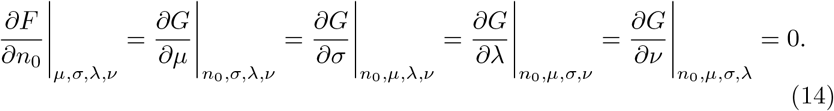

The resulting increase in *χ*^2^ w.r.t. (11), Δ*χ*^2^, is also *χ*^2^ distributed with one degree of freedom, because the extra constraint enforced by *ν* has likewise increased the degrees of freedom by one^3^. Thus, to obtain the 95 % confidence interval in *η*, one needs to find the value of *η*_0_ such that (14) leads to a *χ*^2^ increase of Δ*χ*^2^ = 3.8. This procedure applies if *η* is a mean, variance, or quantile, or any other attribute of the distribution.

By adopting the aforementioned procedure, we obtained the best-fit parameters as shown in Table 1, the goodness of fit in Figures 1 and 2, cumulative frequencies in Figures 3 and 4, and 95 % confidence contours in Figure 5, for the two age groups 15-64 and > 65.

**Table 1:** Parameters of the best log normal fit to the two age groups. *µ* and *σ* are as defined in (3); while the other parameters are calculated by applying the best-fit model to the incubation time range of 17 days, which is the full range spanned by the data (true for both age groups). The information on each parameter comprises an expectation value sandwiched between the lower and upper uncertainty limit, both of which are 95 % confidence (note that for the 65+ age group their upper *µ* and *σ* uncertainties are not constrained by the data, because the data have not revealed the other side of the peak of the differential probability distribution, see Figures 2 and 5).

| Parameter | Age 15-64 | Age 65-86 |
| --- | --- | --- |
| Sample size $N$ | 110 | 22 |
| $\mu$ | 1.95, 2.26, 2.95 | 2.50, 4.10, – |
| $\sigma$ | 0.68, 0.83, 1.20 | 0.55, 1.11, – |
| Mean (geometric) | 6.09, 6.85, 7.62 | 8.57, 10.07, 11.54 |
| Mean (arith.) | 7.32, 8.08, 8.88 | 9.60, 11.05, 12.32 |
| Standard deviation (arith.) | 3.90, 4.22, 4.35 | 3.62, 4.09, 4.80 |
| Median | 6.10, 7.00, 8.10 | 8.9, 10.9, 12.6 |
| 0.05 quantile | 1.5, 1.7, 2.1 | 1.65, 3.40, 7.30 |
| Lower quartile ie | 3.40, 4.10, 4.80 | 4.80, 7.40, 9.30 |
| Upper quartile | 9.5, 10.7, 11.9 | 12.6, 14.0, 15.0 |
| 0.9 quantile | 12.9, 14.1, 14.8 | 15.1, 15.8, 16.3 |
| 0.95 quantile | 14.7, 15.4, 15.9 | 16.3, 16.4, 16.6 |

**Figure 1:**
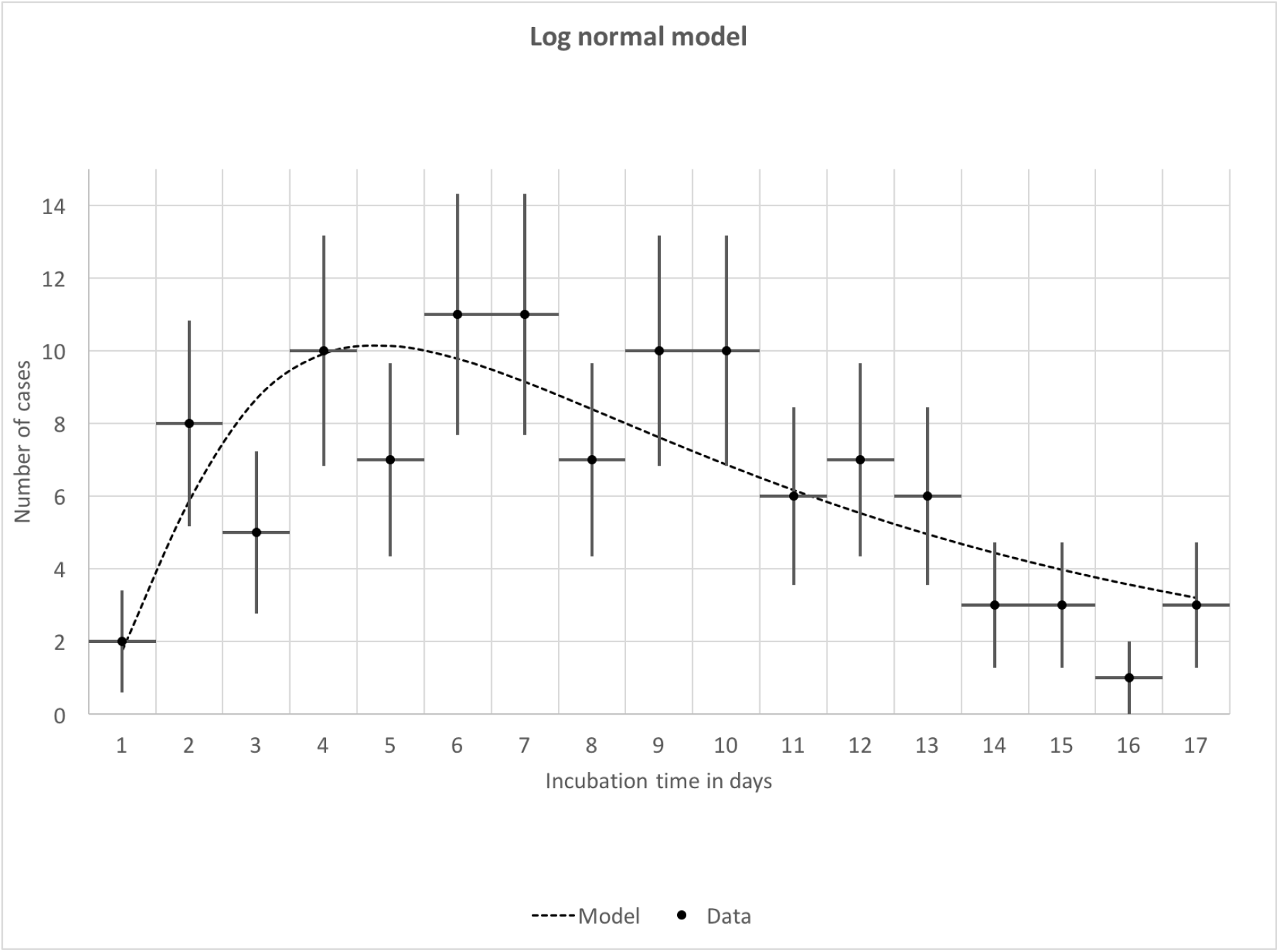
Best fit log normal distribution of the 15-64 age group is plotted against the data. Vertical error bars are the Poisson 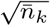 uncertainties in the counts, while horizonal bars mark the duration of 1 day for each time bin. The goodness-of-fit is given by *χ*^2^ = 9.44 for 15 d.o.f., or a 85.3 % probability of rejecting the null hypothesis (namely the hypothesis that the match between data and model occurred purely by random chance).

**Figure 2:**
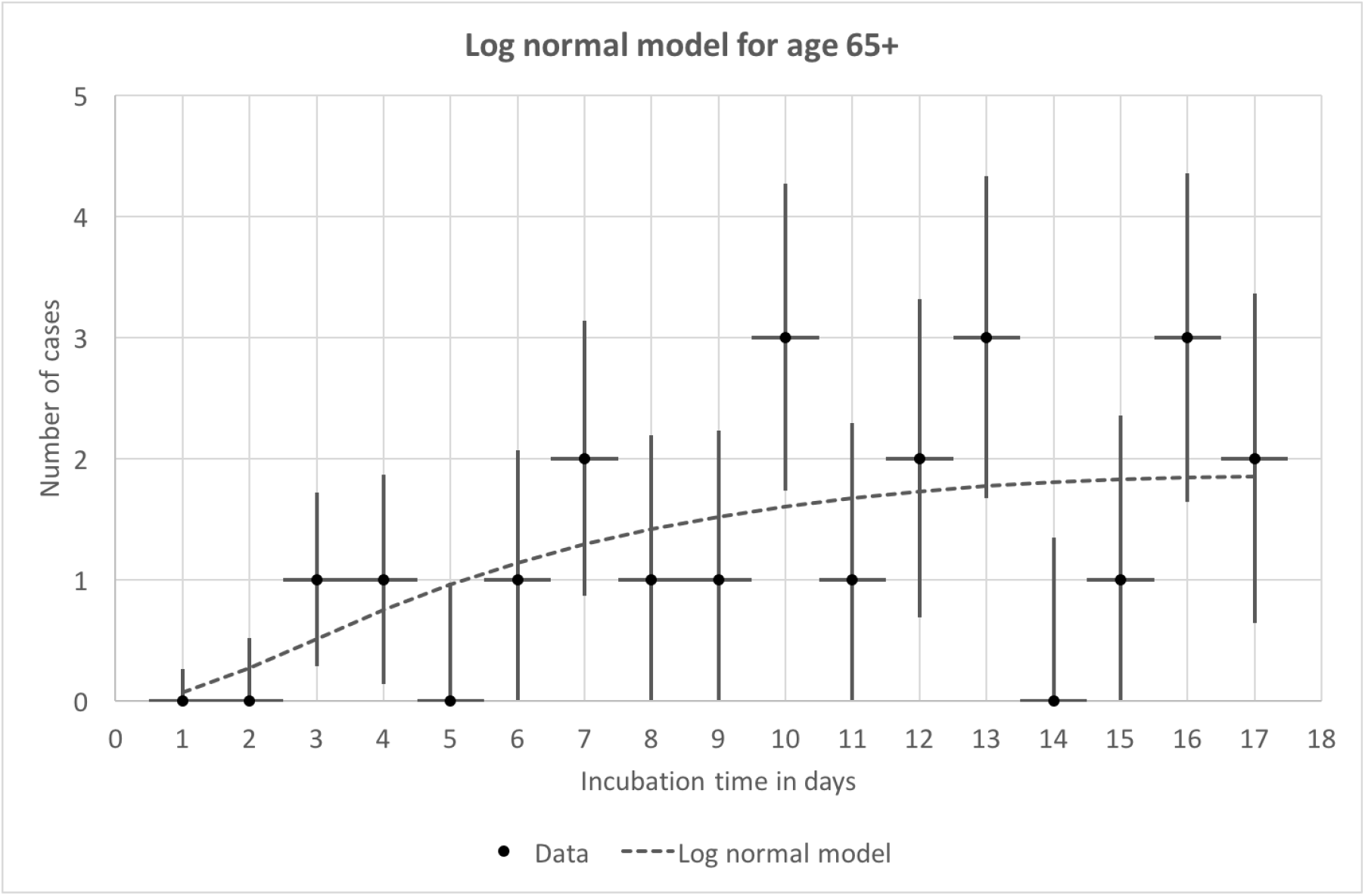
Best fit log normal distribution of the 65-86 age group is plotted against the data. Vertical error bars are the Poisson 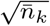 uncertainties in the counts, while horizonal bars mark the duration of 1 day for each time bin. The goodness-of-fit is given by *χ*^2^ = 7.85 for 15 degrees of freedom, or a 93.0 % probability of null hypothesis rejection (for comparison the Gumbel (log Weibull) model scored, under the same fitting criteria, *χ*^2^ = 8.17, or 91.7 % probability of null hypothesis rejection). Note however that for this age group the data have not revealed the other side of the peak of the differential probability distribution.

**Figure 3:**
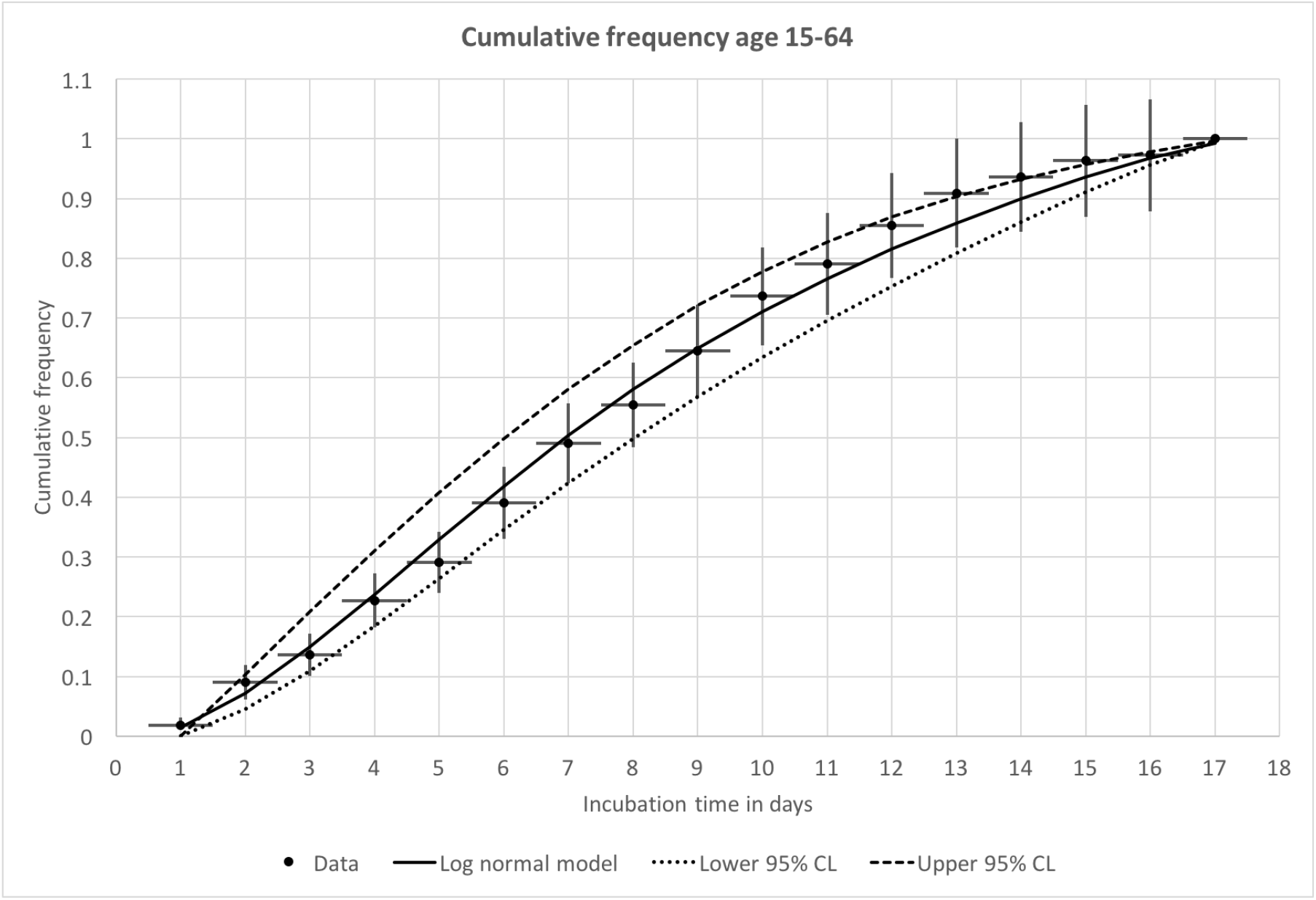
Cumulative frequency distribution for the 15-64 age group. Solid line is the best-fit log normal model for the differential distribution, while dotted lines mark the 95 % confidence (or Δ*χ*^2^ = 3.8 for one degree of freedom, see text) uncertainties of the model. Vertical error bars for the data are Poisson counting fluctuations (1-*σ*, and correlated among the time bins because of the accumulation of counts), while horizonal bars mark the duration of 1 day for each time bin.

**Figure 4:**
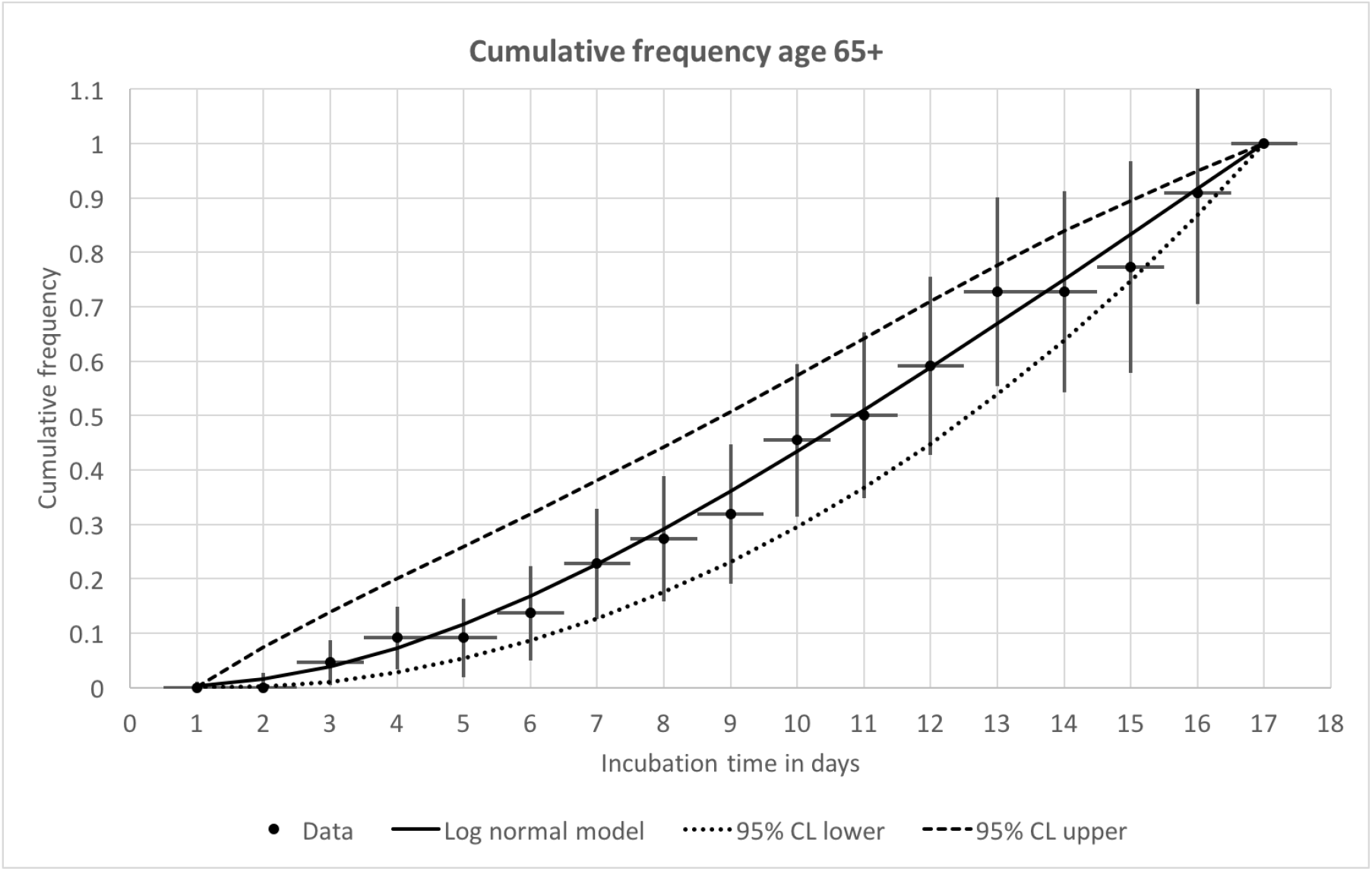
Cumulative frequency distribution for the 15-64 age group. For more information see the caption of the previous graph.

**Figure 5:**
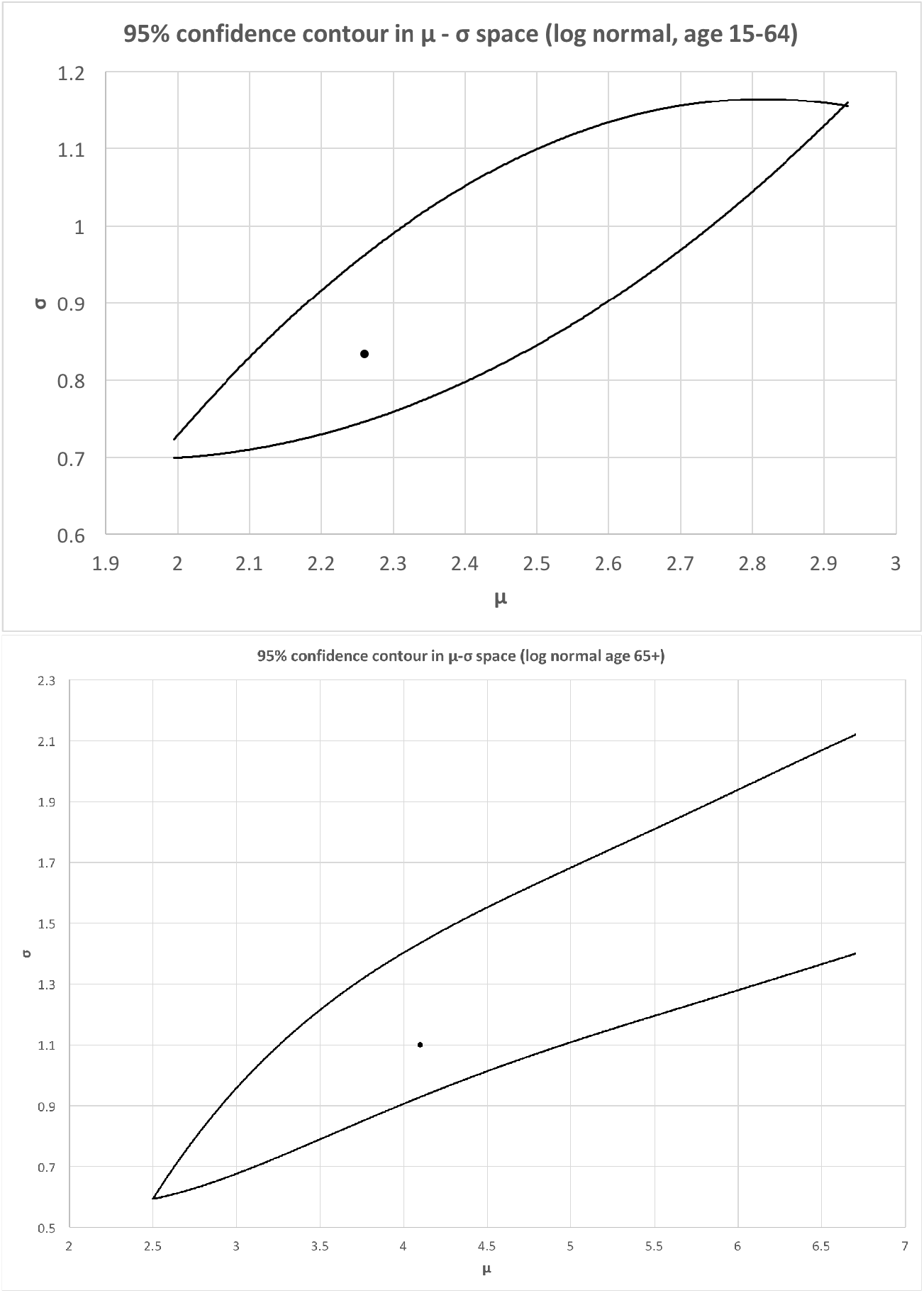
95 % (*i*.*e. χ*^2^ + 3.8) confidence contours identifying the likelihood of pairs of *µ* and *σ* values. The central black dot locates the best fit model. Note the upper limits of both model parameters are unconstrained by the data in the case of the 65-86 age group.

## 4. Conclusion

The two age groups being analyzed are clearly distinct samples. For the 15 - 64 age group, the median incubation time of 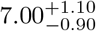 days (uncertainties are 95 % CL, see Table 1) is broadly consistent with previous measurements [2, 3, 4, 5, 6]. For the 65-90 age group, the median is 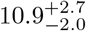 days is statistically significantly longer. The other equally importantly point is, as revealed by the open confidence contour in Figure 5b, the inability of the 65-86 age group data in constraining the upper bound of the model parameters *µ, σ*. This is because the sample of *N* = 22 cases here has a maximum incubation time of 17 days, beyond which one ran out of data even though the differential case histogram has not yet peaked (Figure 3). This indicates the potential of much longer Covid-19 incubation time for age 65-86 years old patients. It should also be pointed out that we attempted another model, *viz*. the Gumbel (or log Weibull) distribution, to see if the situation improves, but the goodness-of-fit turns out to be worse than the log normal (see the caption of Figure 3 for details). Indeed, only a larger sample for this age group can settle the question of the true median incubation time by clinching the tail of the distribution, equivalently by closing the 95 % CL contour of Figure 5b.

## Data Availability

All the data used are available in the public domain

http://wjw.sz.gov.cn/yqxx/

## Footnotes

1 Under this assumption one can also derive (3) without enlisting the Central Limit Theorem, by consideration the viral interaction with the human body as a thermodynamic process with a fixed mean and variance, *i*.*e*. one which maximizes the Entropy − ∑_*j*_ *p*_*j*_ ln *p*_*j*_ subject to the constraints *p*_*j*_ = 1, ∑_*j*_ *p*_*j*_ ln *τ*_*j*_ = *µ*, and ∑_*j*_ *p*_*j*_ (ln *τ*_*j*_)^2^ = *σ*^2^ + *µ*^2^.

2 The maximum likelihood method only works when there is *independence* of measurement intervals. Thus it is *incorrect* to apply the model to fitting the data of a cumulative (or integral) distribution, in which the counts of previous intervals affect the later ones.

3 To elaborate, the degrees of freedom equals the number of independent data points minus the number of free parameters in the model. If the last of the three quantities is reduced by one because of the incorporation of the *ν*-related constraint, the first will be increased by one.

